# Prenatal Lead Exposure is Negatively Associated with the Gut Microbiome in Childhood

**DOI:** 10.1101/2023.05.10.23289802

**Authors:** Shoshannah Eggers, Vishal Midya, Moira Bixby, Chris Gennings, Libni A Torres-Olascoaga, Ryan W. Walker, Robert O. Wright, Manish Arora, Martha María Téllez-Rojo

## Abstract

**Background:** Metal exposures are associated with gut microbiome (GM) composition and function, and exposures early in development may be particularly important. Considering the role of the GM in association with many adverse health outcomes, understanding the relationship between prenatal metal exposures and the GM is critically important. However, there is sparse knowledge of the association between prenatal metal exposure and GM later in childhood.

**Objectives:** This analysis aims to identify associations between prenatal lead (Pb) exposure and GM composition and function in children 9-11 years old.

**Methods:** Data come from the Programming Research in Obesity, Growth, Environment and Social Stressors (PROGRESS) cohort based in Mexico City, Mexico. Prenatal metal concentrations were measured in maternal whole blood drawn during the second and third trimesters of pregnancy. Stool samples collected at 9-11 years old underwent metagenomic sequencing to assess the GM. This analysis uses multiple statistical modeling approaches, including linear regression, permutational analysis of variance, weighted quantile sum regression (WQS), and individual taxa regressions, to estimate the association between maternal blood Pb during pregnancy and multiple aspects of the child GM at 9-11 years old, adjusting for relevant confounders.

**Results:** Of the 123 child participants in this pilot data analysis, 74 were male and 49 were female. Mean prenatal maternal blood Pb was 33.6(SE=2.1) ug/L and 34.9(SE=2.1) ug/L at second and third trimesters, respectively. Analysis suggests a consistent negative relationship between prenatal maternal blood Pb and the GM at age 9-11, including measures of alpha and beta diversity, microbiome mixture analysis, and individual taxa. The WQS analysis showed a negative association between prenatal Pb exposure and the gut microbiome, for both second and third trimester exposures (2Tβ=-0.17,95%CI=[-0.46,0.11]; 3Tβ=-0.17,95%CI=[-0.44,0.10]). Ruminococcus gnavus, Bifidobacterium longum, Alistipes indistinctus, Bacteroides caccae, and Bifidobacterium bifidum all had weights above the importance threshold from 80% or more of the WQS repeated holdouts in association with both second and third trimester Pb exposure.

**Discussion:** Pilot data analysis suggests a negative association between prenatal Pb exposure and the gut microbiome later in childhood; however, additional investigation is needed.

## Introduction

Lead (Pb) has been a recognized environmental hazard for centuries,(Woolley, 1984) however its etiological pathways to disease are still not entirely understood. One potential mechanistic pathway between Pb exposure and its many downstream adverse health outcomes may be via the human gut microbiome. The collection of trillions of microbes that inhabit the human gut, including bacteria, fungi, viruses, and archaea, as well as their many genetic functions, are known as the gut microbiome.(Human Microbiome Project Consortium, 2012) The normal functions of the gut microbiome include nutrient metabolism, support of the mucosal and epithelial barriers within the gut, and interactions with immune, nervous, and cardiovascular systems.(Gomaa, 2020; Wilmes et al., 2022) Xenobiotic exposures, including Pb, influence the composition and function of the gut microbiome, affecting its interaction with systemic bodily function, and may lead to altered health status.(Claus et al., 2016)

Animal studies have linked Pb exposure to alterations of the gut microbiome including shifts in individual sample diversity (α-diversity), community composition (β-diversity), and individual bacterial taxa and gene abundance (Breton et al., 2013; Wu et al., 2016; Gao et al., 2017; Xia et al., 2018). Moreover, the body of epidemiologic evidence linking Pb exposure to altered gut microbiome composition and function is growing rapidly. Studies have linked human Pb exposure to increased abundance of many specific bacteria, including Proteobacteria (Bisanz et al., 2014; Eggers et al., 2019), a common indicator of gut microbiome dysbiosis, or imbalance (Shin et al., 2015; Litvak et al., 2017). Other analyses have linked Pb exposure to bacteria that are known to affect gut barrier integrity and gut health.(Laue et al., 2020; Sitarik et al., 2020; Shen et al., 2022) Alterations in gut and blood-brain barrier integrity resulting from metal-associated shifts in the gut microbiome may allow for increased metal toxicity by allowing for greater absorption into the bloodstream. While this growing field of epidemiologic research has uncovered relationships between Pb exposure and the gut microbiome, so far the analyses have been limited to the use of 16S rRNA amplicon sequencing data, which is less accurate in assigning taxonomy and inferring gene function than metagenomic sequencing. More studies with advanced omics analysis are needed to understand the relationship between Pb exposure and the gut microbiome.

Little is known about the relationship between prenatal exposures and the gut microbiome later in childhood. In fact, there are relatively few studies that investigate the gut microbiome in general between the ages of 5 and 20, especially from populations in low to middle-income countries.(Ortega, 2022) Using data and specimens from the Programming Research in Obesity, Growth, Environment and Social Stressors (PROGRESS) cohort, we examine relationships between prenatal Pb exposure and the gut microbiome of 9-11 year old children from Mexico City, Mexico. In this study, we aim to identify critical windows of prenatal Pb exposure that are associated with the gut microbiome later in childhood. Given the existing evidence from epidemiological and animal studies, we hypothesize that elevated prenatal Pb exposure during at least one time point will be associated with altered gut microbiome composition at 9-11 years old.

## Methods

### Study Design

PROGRESS is an ongoing prospective birth cohort in Mexico City, Mexico. The study enrolled 948 women in early pregnancy who went on to a live birth through the Mexican Social Security System, followed the offspring in infancy every 6 months, and biannually thereafter. The focus of PROGRESS is on neurobehavioral development and child obesity, with emphasis on environmental exposures, like metals, in pregnancy and early life that program later life behavior and growth. Exposures and outcomes were assessed at several time points beginning in the 2^nd^ trimester of pregnancy through a combination of surveys, physical exams, psychological and behavioral assessments. At each visit, biological specimens (including blood) were collected, processed, aliquoted and stored. Stool samples were collected from a subset of participants (n=123) when the children were between the ages of 9-11. Protocols for the main PROGRESS study, and its ancillary microbiome study were reviewed and approved by the Institutional Review Board at the Icahn School of Medicine at Mount Sinai (STUDY-12-00751A, STUDY-21-00242).

### Pb Exposure Measurement

Prenatal Pb exposure in the PROGRESS cohort has been previously described. (Heiss et al., 2020) Briefly, Pb exposure was assessed using maternal whole blood, drawn during the second and third trimesters of pregnancy, at an average of 18.3 and 31.6 weeks gestation, respectively. Pb level was measured using inductively coupled plasma mass spectrometry (ICP-MS) in the trace metals laboratory at the Icahn School of Medicine at Mount Sinai.

### Gut Microbiome Sample Collection and Processing

Participants were recruited at the PROGRESS clinic visit as part of the 9-11 year visit. Samples were collected in clinic or at home using a sampling kit provided. Once obtained, stool samples were stored in a biosafety bag in the participant’s home refrigerator at 4°C. The sample was retrieved by a driver within 24 hours, processed following the FAST(Romano et al., 2018) protocol, and stored at -70°C within 48 hours from sample deposit. Frozen samples were sent to the Microbiome Translational Center at Mount Sinai. Samples were then processed and sequenced in two batches, with n=50 and n=73 samples, respectively. Whole genome sequencing was performed using the NEBNext DNA Library Prep kit and sequenced on an Illumina HiSeq. Sequencing reads were quality trimmed with Trimmomatic (Bolger et al., 2014) and human reads removed by mapping to a reference with bowtie2. (Langmead and Salzberg, 2012) The remaining reads were processed using MetaPhlAn2 (Truong et al., 2015) and StrainPhlAn (Truong et al., 2017) to determine microbial taxonomy down to the species/strain level, and HUMAnN2 (Franzosa et al., 2018) to profile microbial gene pathways.

### Covariates

Covariates used in this analysis included child sex, child’s age at time of stool sample collection, mother’s socio-economic status (SES) during pregnancy, mother’s age at birth, mother’s body mass index (BMI) during pregnancy, and microbiome analysis batch. Mother’s height and weight were collected with a professional digital scale and stadiometer at the study visit during the second trimester of pregnancy. Weight and height were used to calculate BMI, which was modeled as a continuous covariate in regression analyses. SES during pregnancy was assessed based on the 1994 Mexican Association of Intelligence Agencies Market and Opinion (AMAI) rule 13*6. Families are classified in six levels based on 13 questions about characteristics of the household. Most families in the study were low to middle SES, thus the six categories were condensed into three: lower, middle, and higher.(Sanders et al., 2022)

### Statistical Analysis

All analysis was conducted in R version 4.0.3. Any two tailed p-value less than 0.05 was considered statistically significant.

#### Data Processing

The count data for each Taxa were converted into relative abundance. To consider any possible batch effect while measuring taxa count, (1) only those taxa with at least 5% relative abundance in both batches were further considered in primary statistical analyses; (2) All models were controlled for a batch indicator variable along with other covariates. (3) Further detailed sensitivity analysis was conducted by choosing taxa with at least 25% relative abundance in both batches. The second and third-trimester Pb concentrations were log (base = 2) transformed to meet distributional assumptions with higher confidence.

#### Alpha & Beta Diversity

We calculated Shannon alpha diversity and Bray-Curtis beta diversity.(Shannon, 1948; Bray and Curtis, 1957) To estimate the associations with second and third-trimester lead concentrations, (1) for alpha diversity, we used the Kruskal-Wallis rank sum test (without covariate adjustment) and linear regression (with covariate adjustment); and (2) for beta diversity, we used PERMANOVA with 10000 permutations and w/o covariate adjustments.(Oksanen et al., 2019)

#### Weighted Quantile Sum Regression

We used the Weighted Quantile Sum (WQS_RSRH_) regression (Carrico et al., 2015) with random subset (Curtin et al., 2019) and repeated holdouts (Tanner et al., 2019), as established for microbiome data (Eggers et al., 2023), to estimate the association between second and third-trimester lead exposures and the effect on the abundance of the overall mixture of microbial taxa. Since the interest lies in the directionality of the association, the WQS_RSRH_ model was fitted with all the chosen taxa as exposures and the log transformed Pb concentration as the outcome. For ease of interpretation, the relative abundance of the taxa was converted into deciles, while a null relative abundance was kept at zero. Moreover, two WQS_RSRH_ models were fitted at each trimester, with the overall mixture effect assumed in the negative or positive directions. The final optimal model (Liao et al., 2018) from each trimester was chosen based on the smallest Akaike information criterion (AIC) (Akaike, 1974) and the Bayesian information criterion (BIC) (Schwarz, 1978). Those microbial taxa were judged important in the final chosen models, which had weight contributions to the overall mixture index above a chance threshold (1/the number of components in the index).

Lastly, to account for any between taxa correlations and the relatively smaller sample size, each WQS_RSRH_ model was fitted based on 200 repeated holdouts (with randomly 40% data set aside for validation) and 100 bootstrapped with-replacement sampling at each iteration.

#### Taxa-Wide Association Analysis

To estimate the effect of Pb exposure on individual relative abundance of each chosen taxa, we conducted Taxa-wide association analysis (TWAS) with generalized linear models at both trimesters. Further, the unadjusted raw p-values with respect to regression beta estimates were plotted through Volcano plots. The Bonferroni procedure was adapted for multiple comparison error correction on raw p-values. We estimated the effective number of tests for the TWAS using the eigenvalues of the relative abundance of the correlation matrix (Li and Ji, 2005; de Prado-Bert et al., 2021).

#### Gene Function Analysis

We extracted the microbial gene function pathways of the important taxa from the WQS_RSRH_ analysis at each trimester. We selected the top 20 most frequently occurring pathways in each trimester for ease of interpretation. Through a simple Venn diagram, we further elaborated on the pathways which are common to a) both trimesters, b) only present in the second trimester, and lastly, c) only present in the third trimester.

#### Sensitivity Analysis

We also conducted a sensitivity analysis to understand whether the choice of relative abundance in both batches affected the overall mixture effect. To that end, we repeated the entire WQS_RSRH_ analysis with only those taxa having at least 25% relative abundance in both batches. Further, we chose not to rescale the chosen taxa with at least 5% relative abundance (1) to reflect the original contributions and (2) to make the analysis robust irrespective of the chosen relative abundance cutoff.

#### Covariate Adjustment

Each model was controlled by *a priori* chosen set of covariates. Although we thought Pb exposure during childhood may be a confounder, we tested the correlation between prenatal Pb and childhood Pb at birth, 1 year, 2 years, and 4 years of age, and did not find a correlation, thus we decided to exclude these variables from the models. A few covariates had missing values (less than 5%), which were imputed by the multiple imputation chained equations as implemented in the “MICE” R package (Buuren and Groothuis-Oudshoorn, 2011).

## Results

### Study Population Characteristics

Of 123 participants in this study (Table 1), 49 were female, and 74 were male. Mean Pb concentration was 33.6 ug/L and 34.9 ug/L in the second and third trimesters of pregnancy, respectively. Mothers with low SES were more likely to be in the fourth quartile of Pb exposure for both trimesters of pregnancy.

**Table 1.** Characteristics of the analytical study population by Pb exposure in the second and third trimester of pregnancy.

|  | Total | Quartile 1 | Quartile 2 | Quartile 3 | Quartile 4 |
| --- | --- | --- | --- | --- | --- |
|  | N = 123 | Mean (SE) | Mean (SE) | Mean (SE) | Mean (SE) |
| *Exposure* |  | *n (%)* | *n (%)* | *n (%)* | *n (%)* |
| Second Trimester Pb (ug/L) | 33.6 (2.1) | 13.9 (0.5) | 22.2 (0.4) | 32.9 (0.9) | 65.5 (4.3) |
| *Covariates* |  |  |  |  |  |
| Child Sex | 123 |  |  |  |  |
| Male | 74 (60.2) | 10 (58.8) | 19 (59.4) | 19 (65.5) | 26 (57.8) |
| Female | 49 (39.8) | 7 (41.2) | 13 (40.6) | 10 (34.5) | 19 (42.2) |
| Maternal SES |  |  |  |  |  |
| Lower | 66 (53.6) | 7 (41.2) | 16 (50.0) | 16 (55.2) | 27 (60.0) |
| Medium | 45 (36.6) | 8 (47.0) | 12 (37.5) | 11 (37.9) | 14 (31.1) |
| Higher | 12 (9.8) | 2 (11.8) | 4 (12.5) | 2 (6.9) | 4 (8.9) |
| Maternal age at pregnancy (years) | 28.5 (0.5) | 28.9 (0.5) | 27.2 (0.5) | 29.8 (0.6) | 28.3 (0.5) |
| Maternal BMI during pregnancy (kg/m<sup>2</sup>) | 27.2 (0.4) | 26.6 (0.4) | 27.8 (0.4) | 27.1 (0.3) | 27.0 (0.5) |
| Child age at gut microbial sample collection (years) | 9.7 (0.7) | 9.7 (0.1) | 9.5 (0.1) | 9.6 (0.1) | 9.8 (0.1) |
|  | N = 123 | Mean (SE) | Mean (SE) | Mean (SE) | Mean (SE) |
| *Exposure* |  | *n (%)* | *n (%)* | *n (%)* | *n (%)* |
| Third Trimester Pb (ug/L) | 34.9 (2.1) | 14.6 (0.5) | 23.6 (0.5) | 36.4 (1.1) | 66.4 (4.4) |
| *Covariates* |  |  |  |  |  |
| Child Sex | 123 |  |  |  |  |
| Male | 74 (60.2) | 14 (77.8) | 15 (53.6) | 21 (61.8) | 24 (55.8) |
| Female | 49 (39.8) | 4 (22.2) | 13 (46.4) | 13 (38.2) | 19 (44.2) |
| Maternal SES |  |  |  |  |  |
| Lower | 66 (53.6) | 8 (44.4) | 15 (53.6) | 24 (70.6) | 19 (44.2) |
| Medium | 45 (36.6) | 7 (38.9) | 11 (39.3) | 10 (29.4) | 17 (39.5) |
| Higher | 12 (9.8) | 3 (16.7) | 2 (7.1) | 0 (0.0) | 7 (16.3) |
| Maternal age at pregnancy (years) | 28.5 (0.5) | 29.6 (0.5) | 28.4 (0.5) | 27.5 (0.5) | 28.7 (0.5) |
| Maternal BMI during pregnancy (kg/m<sup>2</sup>) | 27.2 (0.4) | 27.5 (0.4) | 26.7 (0.3) | 27.2 (0.5) | 27.4 (0.4) |
| Child age at gut microbial sample collection (years) | 9.7 (0.7) | 9.8 (0.1) | 9.5 (0.1) | 9.86 (0.1) | 9.6 (0.1) |

### Alpha & Beta Diversity

In linear regression analysis of alpha (within individual) diversity and prenatal Pb exposure, we found slight negative associations, that were not statistically significant, for both second and third trimester Pb exposures, in unadjusted and adjusted models (Table 2). In PERMANOVA analysis of beta (between individual) diversity (Table 2), second trimester Pb exposure was associated with a non-significant R^2^ of less than 1% (Adjusted R^2^ = 0.007, p-value = 0.515). However, third trimester Pb exposure was associated with an R^2^ of 1.1% in the adjusted model, and was directionally trending (Adjusted R2 = 0.011, p-value = 0.066).

**Table 2:** Estimates of association between alpha and beta diversity and prenatal Pb exposure.

|  | Alpha Diversity (Shannon)*<br>Beta (p-value) | Beta Diversity (Bray-Curtis)<sup>#</sup><br>R<sup>2</sup> (p-value) |
| --- | --- | --- |
| Second Trimester Pb | -1.48 (0.38) | 0.73 % (0.52) |
| Third Trimester Pb | -1.26 (0.45) | 1.12 % (0.06) |

\*Linear model adjusted for covariates, # PERMANOVA with adjusted covariates

### Microbiome Mixture Analysis

The primary WQS_RSRH_ analysis was run in the negative direction because we hypothesized the association between prenatal Pb exposure and the gut microbiome mixture to be negative (Figure 1). Including adjustment for covariates, second trimester Pb exposure was negatively associated with the gut microbiome mixture (β = -0.17, 95%CI = [-0.46, 0.11]), with 88% of the repeated holdout estimates below zero. Third trimester Pb exposure showed a very similar association with the gut microbiome mixture (β = -0.17, 95%CI = [-0.44, 0.10]), and had 89% of the repeated holdout estimates below zero. Within the weighted indices, taxa with a weight above 0.027 were considered important in the mixture association. Of the 20 bacterial taxa above the importance threshold for second trimester Pb exposure, 16 were also above the importance threshold in association with third trimester Pb exposure. Ruminococcus gnavus, Bifidobacterium longum, Alistipes indistinctus, Bacteroides caccae, and *Bifidobacterium bifidum* all had weights above the importance threshold from 80% or more of the repeated holdouts in association with both second and third trimester Pb exposure.

**Figure 1.**
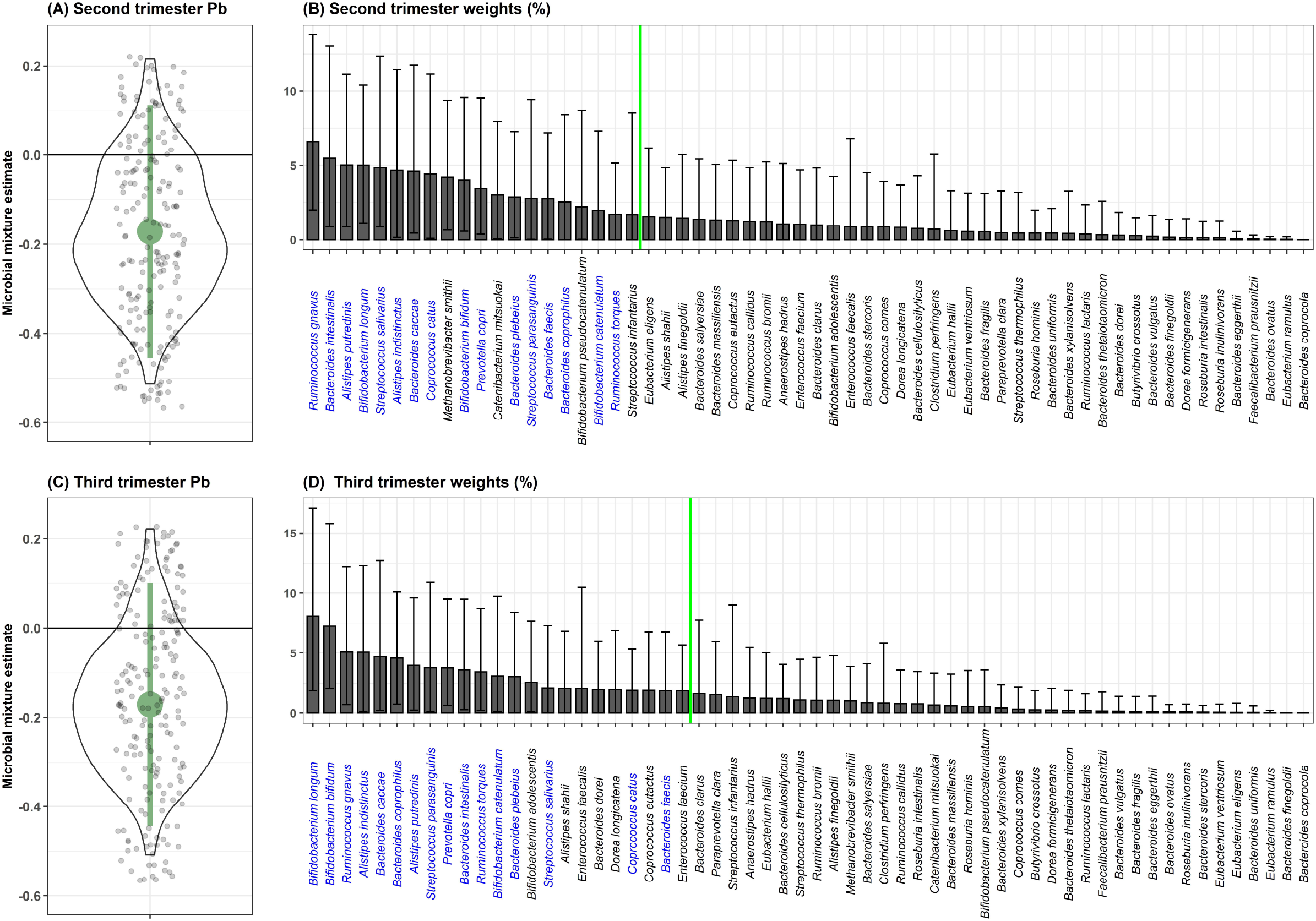
WQS_RSRH_ estimates for the association of the gut microbiome mixture with prenatal Pb exposure in the a) second and c) third trimester of pregnancy. Small grey jitter dots represent the WQS_RS_ estimates from each repeated holdout. The violin plot around those points show the density of holdout estimates at each interval. Avserage percent weight for each taxa within the WQS index are shown for the b) second and d) third trimester Pb exposure. The green line indicates the importance threshold for weights above random chance. Taxa that are labeled in blue are above the threshold for both trimesters of exposure.

In a sensitivity analysis, we ran the same WQS model using only the bacterial taxa that were present in at least 25% (instead of 5%) of participants from both analytical batches, and found associations in the same direction, with slightly larger estimates and confidence intervals that still crossed zero (Sup. Figure 1). We conducted an additional sensitivity analysis running the WQS_RSRH_ analysis in the positive direction and found null results. When comparing the likelihood estimates between the negative and positive WQS_RSRH_ models, the likelihood was higher for the negative model, confirming our appropriate use of the negative model as the primary analysis.

When examining microbial gene function pathways of the important taxa associated with each of the second and third trimester Pb exposure separately, of the top 20 most abundant gene pathways for each trimester, approximately 1/2 of pathways from each trimester were found to be unique (Figure 2). Overall, common pathways were more likely to do with nucleic acid biosynthesis, and functions essential to all bacteria, while pathways associated with only one trimester Pb exposure or the other, were more likely to be involved in amino acid biosynthesis, and more specialized metabolic functions.

**Figure 2.**
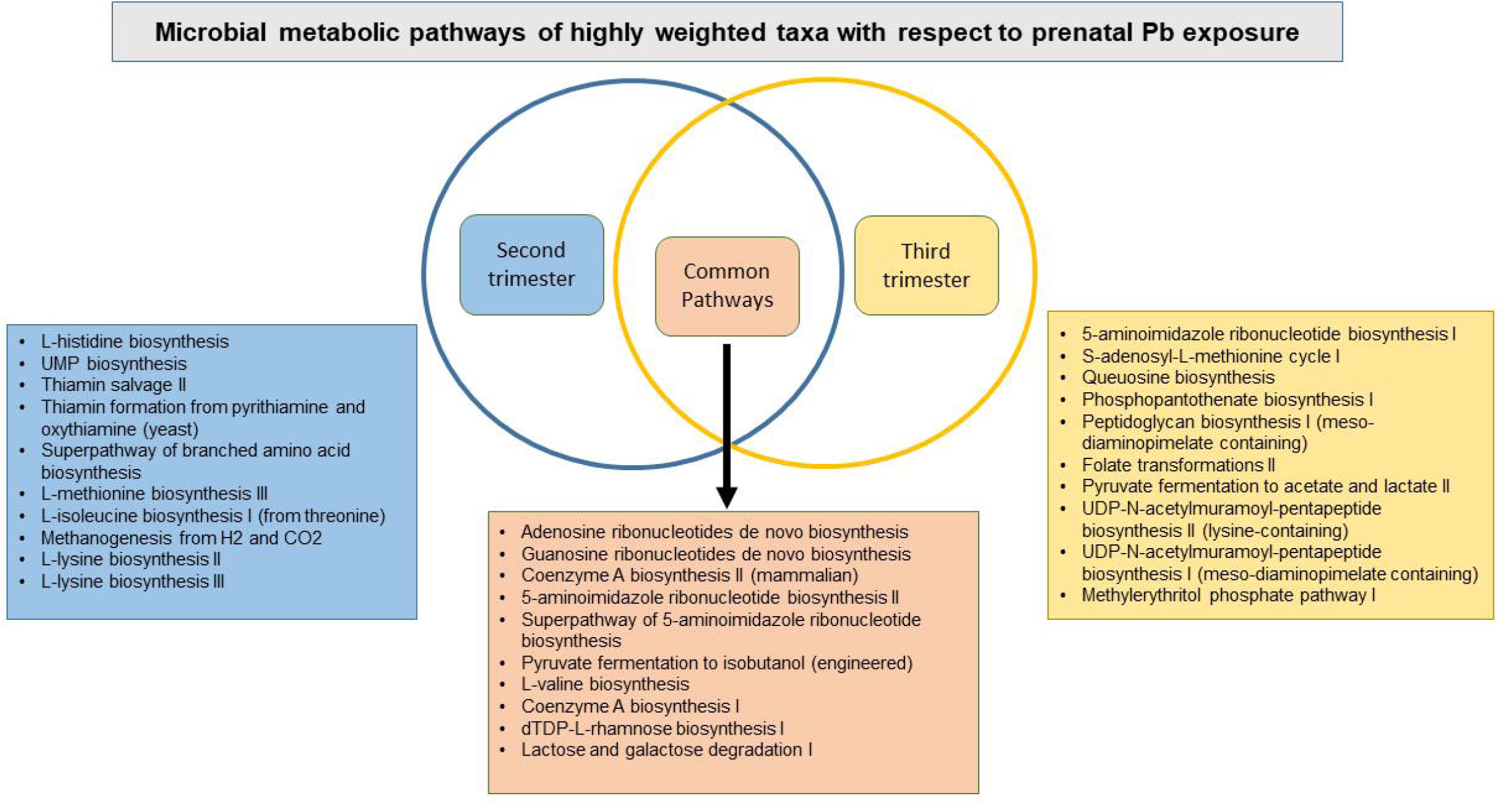
Venn diagram of the top 20 most abundant microbial gene functions from the highly weighted taxa in the WQS_RSRH_ analysis.

### Individual Taxa Analysis

In analysis of each individual bacterial taxa with prenatal Pb exposure (Figure 3), we found six taxa (*Alistipes putredinis, Ruminococcus ghavus, Bacteroides caccae, B. intestinalis, Coprococcus catus*, and *A. indistinctus*) to be negatively associated, and one (*B. coprocola*) to be positively associated with second trimester exposure. With third trimester exposure, we found three taxa (*Bifidobacterium bifidum, B. longum, A. indistinctus*) to be negatively associated, and three taxa (*B. coprocola, Eubacterium eligens, B. finegoldii*) to be positively associated with Pb.

**Figure 3.**
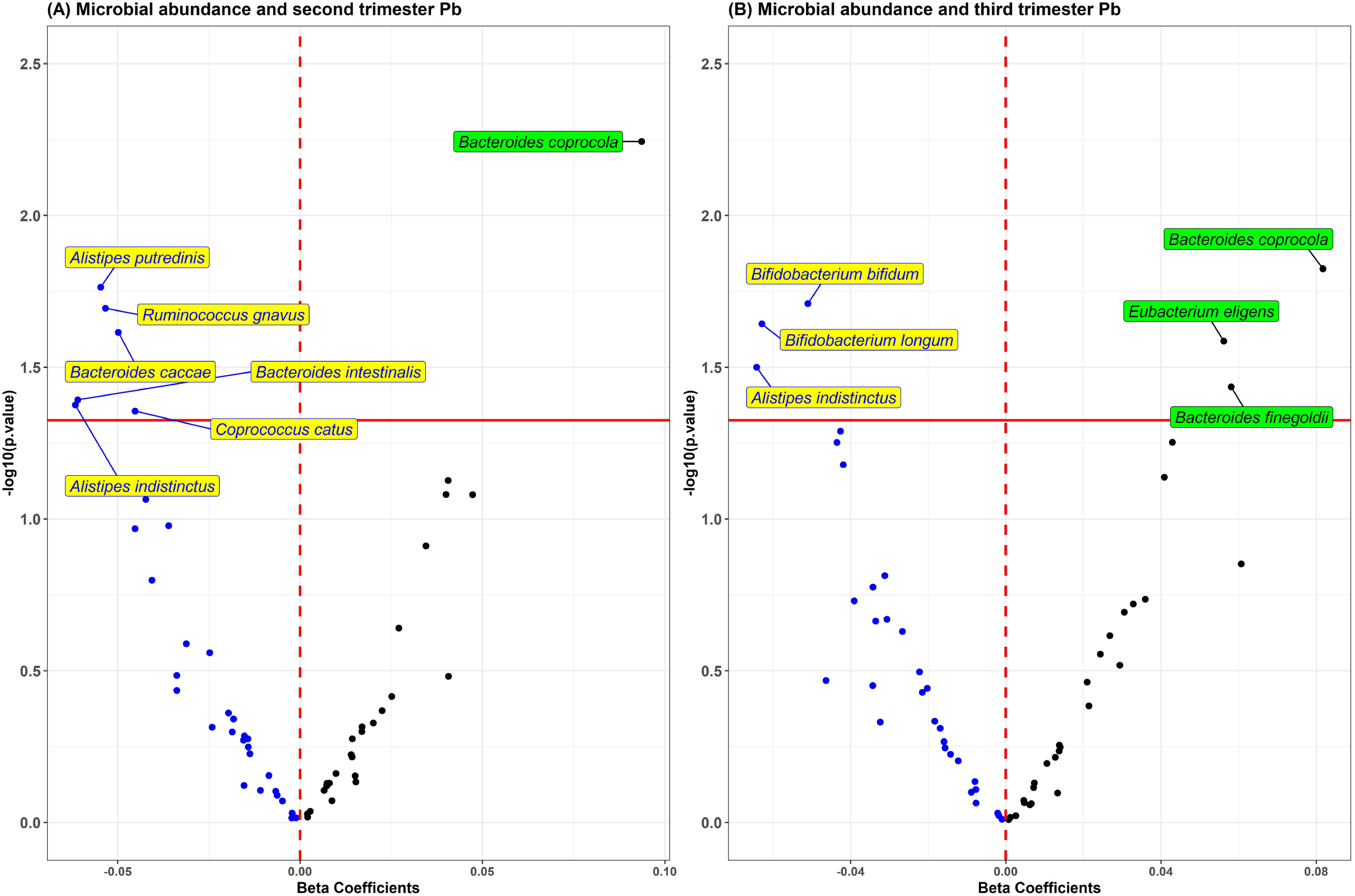
Volcano plot of estimates and p-values from the taxa wide association analysis (TWAS) of bacterial abundance in association with a) second and b) third trimester Pb exposure. Blue dots are taxa associated in the negative direction and black dots are associated in the positive direction.

## Discussion

In this analysis of pilot microbiome data from the PROGRESS cohort, we examined associations between prenatal Pb exposure and several different components of gut microbiome composition and function. We found consistent negative associations between Pb exposure during the second and third trimester of pregnancy and several assessments of gut microbiome composition and function at 9-11 years old. Associations between prenatal Pb and the gut microbiome later in childhood tended to be strongest for second trimester exposure, providing support for the second trimester of pregnancy in particular as a critical window of exposure.

The focus of this analysis was on the association between prenatal Pb exposure and the gut microbiome later in childhood. There are many other potential exposures that may influence the composition and function of the gut microbiome between the prenatal exposure and the gut microbiome at 9-11 years, including continued exposure to Pb, diet, antibiotic use, animal exposure, child BMI and many others. However, none of these factors were included in this analysis because they are not confounders, as they are either 1) not associated with prenatal Pb exposure, or 2) potentially on the causal pathway between prenatal Pb exposure and the gut microbiome later in life.

The underlying mechanism of association between prenatal Pb exposure and the gut microbiome later in childhood could work in multiple ways. Because the gut microbiome is shaped in part by the host immune system, (Petersen et al., 2019; Wong et al., 2022) prenatal Pb exposure may alter the trajectory of immune system development, which can influence gut microbial composition through the life-course. Another possible mechanism is via the maternal gut microbiome. The infant gut microbiome is primarily colonized by bacteria that are transferred from the mother at birth. (Ferretti et al., 2018) Pb exposure in adults also influences gut microbiome composition,(Eggers et al., 2019) thus Pb induced changes in the maternal gut microbiome during pregnancy may be transferred to children at birth, and carried on into childhood, and even later in life.(Gschwendtner et al., 2019) Another potential mechanism may be that maternal Pb exposure is transferred to children through breastmilk, an established Pb exposure pathway (Klein et al., 2017). In that case, postnatal Pb exposure through breastmilk would be along the causal pathway between prenatal Pb exposure and the gut microbiome. Furthermore, all of these mechanisms may be working together. Further mechanistic studies are needed to better understand this complex relationship.

In previously published analyses of prenatal Pb exposure and the gut microbiome in studies with relatively small sample sizes, other researchers have seen similar results. In analysis of a birth cohort based in Detroit, MI (n=146), Sitarik et al, identified associations between prenatal Pb exposure, measured in baby teeth, and decreased abundance of several species of *Bacteroides* within the gut microbiome at 1 month and 6 months of age.(Sitarik et al., 2020) They identified *B. caccae* as being negatively associated with second trimester Pb exposure specifically. We also identified a decreased abundance of *B. caccae* in association with Pb exposure in the second trimester using TWAS, and *B. caccae* was also heavily weighted in WQS_RSRH_ for both trimesters of prenatal Pb exposure. *B. caccae* are common fiber degraders in the human gut microbiome, with mixed health effects. (Wexler, 2007; Chen et al., 2021; Yang et al., 2022; Zhang et al., 2022) This consistent association of prenatal Pb exposure and decreased *B. caccae* abundance both early and late in childhood, from different populations in different countries, is strong evidence of association in this relatively new field.

In another epidemiologic study using data from a Canadian cohort (n=70), Shen et al, found that prenatal Pb exposure, measured in maternal blood, was associated with increased abundance of *Fusobacteriota* in the gut microbiome at 6-7 years of age.(Shen et al., 2022) They did not find a significant association between prenatal Pb and alpha or beta diversity. While the associated gut microbes identified in this analysis and the study by Shen et al, were not the same, it is important to note that links between prenatal Pb and the gut microbiome many years later in childhood were identified, even in these two small data sets. This suggests that there are likely true underlying links between prenatal Pb exposure and the gut microbiome in childhood. Differences in bacterial taxa associated may be due to differences in the study participants, or differences in sample and data processing procedures.

Of all the taxa in this analysis, *Alistipes indistinctus* was the only one identified by both WQS_RSRH_ and TWAS to have a negative association with Pb exposure in the second and third trimesters. *A. indistinctus* is a common member of the human gut microbiome, and relatively newly identified.(Nagai et al., 2010) Little is known about the health implications of *A. indistinctus*. However, one study has identified it as protective against liver fibrosis.(Shao et al., 2018), and another identified it as a keystone species for restoring a healthy gut microbiome in patients with non-alcoholic fatty liver disease.(Wu et al., 2022) Overall, the *Alistipes* genus has been shown to have both beneficial and detrimental health effects in humans.(Parker et al., 2020) Continued investigation of the health implications of *A. indistinctus* are necessary to understand the links between prenatal Pb exposure and downstream health status via the microbiome.

In the analysis of microbial gene pathways from the most highly weighted taxa in the WQS_RSRH_ analysis, approximately 1/3 of the gene pathways were shared between the taxa associated with second and third trimester Pb exposure. With a few exceptions, the pathways that were common among these bacteria are used for nucleic acid biosynthesis and other essential functions for bacterial life.(Tsuchiya et al., 2018) The pathways that were not shared were more likely to be used in amino acid biosynthesis, fermentation, and other metabolic pathways. Because these more varied pathways provide a wider range of functional capabilities, they are more likely to influence host health, although their direct influence is not yet fully understood.

While this study added to the growing field of evidence around the negative relationship between metal exposure and the human gut microbiome, there are some limitations to consider. Because this was a pilot study, sample size was limited, which limited our power to detect associations. However, because results were primarily in the negative direction across multiple analytical approaches, and over 85% of the beta estimates from the WQS repeated holdouts were negative, we have increased confidence in reporting a negative result. Samples from this pilot study were also analyzed in two batches. Our strategy for reducing batch effects limited the breadth of microbiome data we could include in the analysis; however, similar prevalence thresholds are frequently used as data reduction steps in microbiome data analysis. Another statistical limitation was the use of prenatal Pb exposure as an outcome in the WQS_RSRH_ models rather than a predictor. This model structure was necessary due to WQS format. The implications of this limitation are minimal however, as this analysis was used to determine association, not causation. Lastly, the use of maternal blood Pb during pregnancy to measure prenatal Pb exposure is not ideal, as it is not a direct measure of fetal Pb exposure.

In future analyses based off this work, we hope to examine additional and more nuanced relationships between prenatal environmental metal exposures and the gut microbiome in childhood. We plan to do additional sample collection in this cohort that will expand the sample size for future analyses. Furthermore, we are developing additional methodological approaches to examine relationships between chemical exposures and the gut microbiome, and their combined effect on downstream health.

In conclusion, this pilot study found a consistent negative association between prenatal Pb exposure and the gut microbiome in late childhood. These results support a growing body of evidence that human Pb exposure may alter gut microbial composition and function, leading to downstream health implications. More studies with larger sample sizes are needed to better understand this relationship.

## Data Availability

The data used in this study can be made accessible to researchers upon appropriate request with the following restrictions to ensure the privacy of human subjects. Note that access to the data is limited due to a data-sharing agreement approved by the IRB at Mount Sinai. Researchers interested in accessing PROGRESS data must send their resume/CV and CITI training certificates to the IRB chair, Ilene Wilets. They must also submit a data analysis plan to the Principal Investigators for PROGRESS; Robert O. Wright, Martha Tellez-Rojo, and Andrea Baccarelli. Once this process is completed, the PROGRESS data analyst, Nia McRae, will send a de-identified dataset via Box, a secure data-sharing platform.

## 3 Conflict of Interest

MA is an employee and equity holder of Linus Biotechnology Inc., a start-up company of Mount Sinai Health System. The company develops tools for the detection of autism spectrum disorder and related conditions. The following authors report no competing interests: SE, VM, CG, MB, LT, RWW, ROW, MT.

## 4 Author Contributions

SE, VM, CG and MA contributed to conception and design of the study. LT, MT, and ROW contributed to data acquisition. VM and MB performed statistical analysis. SE, VM, CG, and RWW contributed to data interpretation. SE wrote the first draft of the manuscript. VM wrote sections of the manuscript. All authors contributed to manuscript revision, read, and approved the submitted version.

## 5 Funding

This work was supported by the National Institute of Environmental Health Sciences (K99ES032884, P30ES023515, R01ES013744).

## 6 Acknowledgments

The authors would like to acknowledge the entire PROGRESS study team, as well as the participants. We would also like to thank Dr. Jeremiah Faith and the Microbiome Translational Center at the Icahn School of Medicine at Mount Sinai.

## 17 Data Availability Statement

The data that was used in this study can be made accessible to researchers upon appropriate request with the following restrictions to ensure the privacy of human subjects. Note that access to the data is limited due to a data sharing agreement approved by the IRB at Mount Sinai. Researchers that are interested in accessing PROGRESS data must send their resume/CV as well as CITI training certificates to the IRB chair, Ilene Wilets. They must also submit a data analysis plan to the Principal Investigators for PROGRESS; Robert O. Wright, Martha Tellez-Rojo, and Andrea Baccarelli. Once this process is completed, the PROGRESS data analyst, Nia McRae will send a de-identified dataset via Box, a secure data sharing platform.

## Notes

### Author Declarations

The ethics committee/IRB of Icahn School of Medicine at Mount Sinai gave ethical approval for this work.

