## Supplemental Information for "Prenatal Lead Exposure is Negatively Associated with the Gut Microbiome in Childhood"

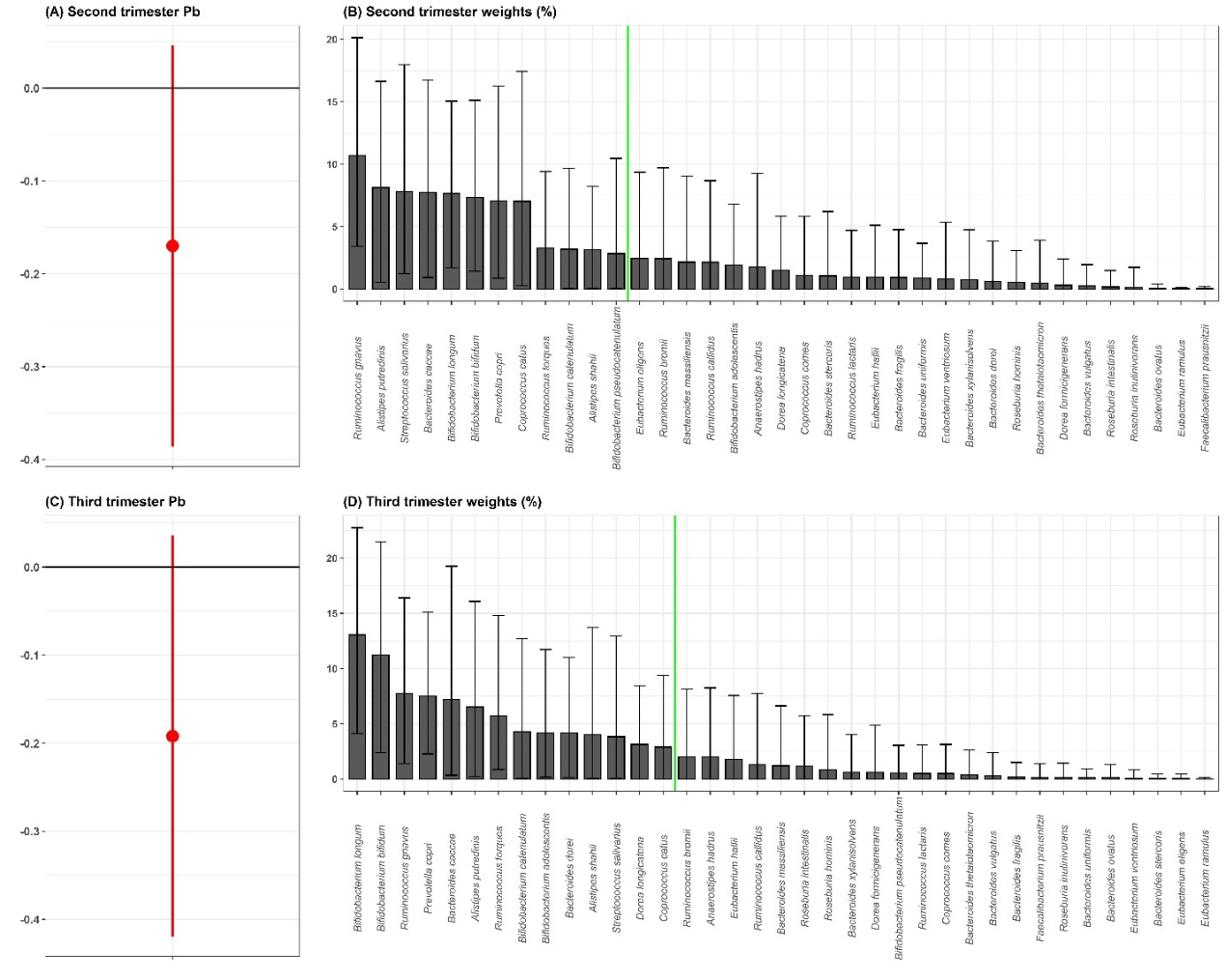

**Supplemental Figure 1.** Sensitivity analysis WQS_RSRH_ estimates for the association of the gut microbiome mixture with prenatal Pb exposure in the a) second and c) third trimester of pregnancy, including only the taxa that are present in at least 25% of participants from each analytical batch. Average percent weight for each taxa within the WQS index are shown for the b) second and d) third trimester Pb exposure. The green line indicates the importance threshold for weights above random chance.

| **Second trimester Pb** | **Third trimester Pb** |
| --- | --- |
| PWY-7219: adenosine ribonucleotides de novo biosynthesis | PWY-7219: adenosine ribonucleotides de novo biosynthesis |
| PWY-7221: guanosine ribonucleotides de novo biosynthesis | COA-PWY-1: coenzyme A biosynthesis II (mammalian) |
| COA-PWY-1: coenzyme A biosynthesis II (mammalian) | PWY-7221: guanosine ribonucleotides de novo biosynthesis |
| HISTSYN-PWY: L-histidine biosynthesis | PWY-6122: 5-aminoimidazole ribonucleotide biosynthesis II |
| PWY-5686: UMP biosynthesis | PWY-6277: superpathway of 5-aminoimidazole ribonucleotide biosynthesis |
| PWY-6122: 5-aminoimidazole ribonucleotide biosynthesis II | PWY-6121: 5-aminoimidazole ribonucleotide biosynthesis I |
| PWY-6277: superpathway of 5-aminoimidazole ribonucleotide biosynthesis | PWY-6151: S-adenosyl-L-methionine cycle I |
| PWY-6897: thiamin salvage II | PWY-6700: queuosine biosynthesis |
| PWY-7111: pyruvate fermentation to isobutanol (engineered) | PANTO-PWY: phosphopantothenate biosynthesis I |
| PWY-7357: thiamin formation from pyrithiamine and oxythiamine (yeast) | PEPTIDOGLYCANSYN-PWY: peptidoglycan biosynthesis I (meso-diaminopimelate containing) |
| VALSYN-PWY: L-valine biosynthesis | PWY-3841: folate transformations II |
| BRANCHED-CHAIN-AA-SYN-PWY: superpathway of branched amino acid biosynthesis | PWY-5100: pyruvate fermentation to acetate and lactate II |
| COA-PWY: coenzyme A biosynthesis I | PWY-6386: UDP-N-acetylmuramoyl-pentapeptide biosynthesis II (lysine-containing) |
| DTDPRHAMSYN-PWY: dTDP-L-rhamnose biosynthesis I | PWY-6387: UDP-N-acetylmuramoyl-pentapeptide biosynthesis I (meso-diaminopimelate containing) |
| HSERMETANA-PWY: L-methionine biosynthesis III | PWY-7111: pyruvate fermentation to isobutanol (engineered) |
| ILEUSYN-PWY: L-isoleucine biosynthesis I (from threonine) | VALSYN-PWY: L-valine biosynthesis |
| LACTOSECAT-PWY: lactose and galactose degradation I | COA-PWY: coenzyme A biosynthesis I |
| METHANOGENESIS-PWY: methanogenesis from H2 and CO2 | DTDPRHAMSYN-PWY: dTDP-L-rhamnose biosynthesis I |
| PWY-2941: L-lysine biosynthesis II | LACTOSECAT-PWY: lactose and galactose degradation I |
| PWY-2942: L-lysine biosynthesis III | NONMEVIPP-PWY: methylerythritol phosphate pathway I |

**Supplemental Table 1:** Top 20 microbial metabolic pathways of highly weighted taxa by trimester of prenatal Pb exposure.

| **Supplemental Table 2.** The 13 variables included in the derivation of the socio-economic status (SES) variable used in this analysis. | |
| --- | --- |
| **Variable** | **Label** |
| Bathrooms | Number of bathrooms with shower |
| Boiler | Do you have a boiler? 0=No, 1=Yes |
| Car | Number of cars |
| Children | Number of children |
| Comp | Do you have a personal computer? 0=No, 1=Yes |
| Floor | Type of flooring: 0=Ground or cement, 1=Other material |
| Lightbulbs | Number of lightbulbs: 1= 5 or less, 2= 6-10, 3= 11-15, 4=16-20, 5= 21 or more |
| Micro | Do you have a microwave? 0=No, 1=Yes |
| Player | Do you have a video or DVD player? 0=No, 1=Yes |
| Rooms | Number of rooms |
| Toast | Do you have a toaster? 0=No, 1=Yes |
| Vacuum | Do you have a vacuum cleaner? 0=No, 1=Yes |
| Washer | Do you have a washing machine? 0=No, 1=Yes |
